# The Helping Older People Engage (HOPE) Study: Protocol & COVID Modifications for a Randomized Trial

**DOI:** 10.1101/2022.04.25.22274283

**Authors:** Kimberly A. Van Orden, Yeates Conwell, Benjamin P. Chapman, April Buttaccio, Alexandra VanBergen, Ellen Beckwith, Angela Santee, Jody Rowe, Deborah Palumbos, Geoffrey Williams, Susan Messing, Silvia Sörensen, Xin Tu

## Abstract

**Objectives:** Evidence-based strategies to reduce loneliness in later life are needed because loneliness impacts all domains of health, functioning, and quality of life. Volunteering is a promising strategy, as a large literature of observational studies documents associations between volunteering and better health and well-being. However, relatively few studies have used randomized controlled trials (RCTs) to examine benefits of volunteering, and none have examined loneliness. The primary objective of the Helping Older People Engage (HOPE) study is to examine the social-emotional benefits of a social volunteering program for lonely older adults. This manuscript describes the rationale and design of the trial.

**Design:** Randomized controlled trial examining an existing community volunteering program available nationwide. We are randomly assigning adults aged 60 or older (up to 300) who report loneliness to 12 months of either AmeriCorps Seniors volunteering program or an active control (self-guided life review). Co-primary outcomes are assessed via self-report—loneliness (UCLA Loneliness Scale) and quality of life (WHOQOL-Bref). This trial is registered at clinicaltrials.gov (NCT03343483)

**Setting:** Lifespan of Greater Rochester, a non-medical, community-based aging services agency (volunteering) and participants’ homes (control).

**Participants:** Adult ages ≥60 years who endorsed feelings of loneliness (score of 6 or greater on the 3-item UCLA Loneliness Scale).

**Intervention:** The interventions are 12-months of volunteer service with the AmeriCorps Seniors program or self-guided life review writing (active control).

**Measurements:** Data were collected at baseline, 3-, 6-, and 12-months (loneliness) and baseline and 12-months (quality of life).

**Timeline:** Enrollment for this trial is underway and expected to finish by May 2022, with completion of follow-up assessments through April, 2023, and completion of primary outcomes soon thereafter.

---

Older adults who are lonely carry increased risk for reduced quality of life,^1^ morbidity,^2-5 6-16^ and mortality.^2,7,8,17,18^ The risk of premature mortality related to loneliness is at least as large as the risks arising from such factors as obesity, physical inactivity, alcohol misuse, and smoking.^17^ While not the norm in later life, a significant portion of older adults experience loneliness^8,18-25^ and its health impacts may be more pronounced in later life.^26^ Loneliness is both an indicator (or component) of healthy aging and a determinant of (or contributor to) healthy aging. Theory and empirical data indicate that humans have an innate ‘need to belong’ to social relationships and groups.^27,28^ When this need is not met, loneliness—the perception of social isolation—emerges.^29^ Research with older adults has confirmed that loneliness is due in part to objective circumstances—increasing disability and frailty, environmental barriers to socialization, and bereavement,^9,30,31^ while other research emphasizes the role of subjective perceptions, such as thinking of oneself as useless,^32^ in causing and perpetuating loneliness.^28,33^

Developing evidence-based strategies to reduce loneliness may be a particularly potent strategy to promote well-being and functioning for older adults, as it impacts all domains of health, functioning, and quality of life.^34^ Meta-analytic reviews indicate that loneliness is responsive to behavioral interventions (reviews included adults of all ages).^35,36^ However, the evidence-base to support recommending one program over another is limited by an absence of replication; lack of clarity on mechanisms and essential intervention components (e.g., group delivery); low generalizability (e.g., enrolling only socially connected participants); and limited engagement in programs outside of research studies.^37^ Most studies examine programs developed specifically for research, while few studies test existing community programs; thus, there is very little data supporting effectiveness of routinely recommended programs (e.g., senior centers, friendly calling). These programs are likely effective for some older adults, to some degree, but providing social contact does not necessarily address loneliness in a meaningful way for all older adults.^37^ Finally, the literature is limited by a lack of attention to providing, rather than receiving support, as a more potent means of improving connectedness in later life.^38^

## Volunteering is a promising strategy to reduce loneliness

Older adults are typically motivated to maintain balanced social relationships that are characterized by reciprocity.^39^ Relationships in later life have been found to terminate when an older adult believes he/she is receiving more support than he/she is providing in the relationship.^40^ Providing, compared to receiving, support, has been shown to be more strongly associated with well-being in later life.^38^ Thus, for older adults seeking to reduce feelings of loneliness, helping others by volunteering may be a potent strategy.^41^ Observational studies have documented numerous associated benefits of volunteering for older adults, including reduced depressive symptoms^42-50^ (as well as reduced stress,^51^ improved perceived mental health^52^ and increased self-efficacy^44^) reduced functional impairment,^45,47,48,53^ reduced physical pain,^54^ increased self-perceived health,^45-48,53,55,56^ increased well-being and life satisfaction,^50,56-61^ increased social connections (including social support,^61,62^ social engagement,^55,63,64^ meeting new people,^57,65^ and number of social ties^46,65^), increased perceptions of usefulness,^43,66^ and reduced/delayed mortality.^45-48,55,63,67-70^

While a large literature of observational studies documents associations between volunteering and better health and well-being, relatively few studies have used randomized controlled trials (RCTs) to examine benefits of volunteering (and rule out confounding or reverse causality). Two published RCTs of volunteering in later life examined the Experience Corps program—a nationwide intensive volunteering experience (15+ hours per week for one to two years) for older adults who are placed as volunteers in city schools to help elementary school children with academic and behavioral skills. In the first trial, results at 4-8 month follow-up indicated greater physical activity, physical strength, social support (people one could turn to for help), and cognitive functioning compared to controls.^62^ The second, larger RCT focused on the primary outcomes of decreased disability in mobility,^71^ increased physical activity,^72^ and prevention of atrophy in brain volume,^73^ all with positive findings. The authors also found that physical, social, and cognitive activity increased due to the program, which suggests intensive volunteering may promote health in later life through increased activity.^74,75^ A third trial of a different intervention examined benefits of a brief behavioral intervention designed to increase motivation for volunteering in older adults and found benefit regarding reduced depressive symptoms for those subjects who increased their volunteering hours.^76^ None of these trials examined loneliness as an outcome or required that subjects report loneliness at baseline.

## The HOPE Study

The primary objective of the Helping Older People Engage (HOPE) study is to examine the social-emotional benefits of a social volunteering program for lonely older adults. The volunteering program tested in our study is the Retired Senior Volunteer Program (RSVP), which is one component of AmeriCorps Seniors, a national program administered by AmeriCorps that links adults 55 years and older to volunteer opportunities in their communities in a wide range of roles that take best advantage of their skills and experience. The U.S. Aging Services Network (ASN) is a national network of community-based social service agencies overseen by the Administration on Aging and includes State Agencies on Aging, Area Agencies on Aging, and Title VI Native American aging programs.^19^ Area Agencies on Aging commonly provide local support to administer AmeriCorps Seniors programs. For the HOPE study, the volunteering intervention involves participation in the RSVP Program overseen by Lifespan of Greater Rochester, which provides aging services to older adults in the Monroe County, NY region.^23^ We selected this volunteering program due to its availability nationwide, thus promoting scalability; its focus on matching older adults to valued activities that match their skill set, promoting acceptability and engagement; and its emphasis on supporting volunteers through regular contact with the volunteer coordinator to navigate problems and provide standardized training, all promoting motivation to start and sustain volunteering.

Employing an existing community program available nationwide and an RCT design with an active control condition, HOPE is a Stage III efficacy design within the *NIH Stage Model for Intervention Development*. ^77^ Given promising findings, it will lead to study of implementation/dissemination and pragmatic trial designs to move rapidly move intervention science into practice. We are randomly assigning adults aged 60 or older (up to 300) who report loneliness to 12 months of either AmeriCorps Seniors volunteering program or an active control (self-guided life review). Life review is a reasonable active control because it is intellectually stimulating (also true of volunteering) but its social component is negligible (unlike volunteering).

## Study aims

Our first aim is to test whether one-year of AmeriCorps Senior volunteering results in reduced loneliness (UCLA Loneliness Scale Version 3) and increased health-related quality of life (WHOQOL-Bref global score) compared to an active control condition (self-guided life review writing). We will also explore whether health-related quality of life domains of social and emotional quality of life demonstrate the greatest improvement in response to volunteering (compared to physical and environmental domains of quality of life).

Our second aim examines mechanisms whereby volunteering may reduce loneliness. Based on prior observational studies of volunteering, we hypothesize that increased purpose in life and increased social engagement (satisfaction with social activities) will account for reductions in loneliness.

Our third aim examines conditions under which volunteering is most effective at reducing loneliness, including ‘dose’ of volunteering (number of hours volunteered) and satisfaction with volunteer placements. Given that functional impairment impacts all proposed mechanisms, we will explore whether benefits of volunteering on loneliness are greatest for those with less functional impairment at baseline. We will also conduct exploratory analyses to examine sexual and gender minority (SGM; e.g., lesbian, gay, bisexual, transgender) identity as a potential moderator of primary outcomes, given significant health disparities faced by this group^78^ and the potential for volunteering to function differently in this sub-group of lonely older adults (e.g., due to experiences such as concealing one’s SGM identity).^79^ We will assess SGM identity as well as one indicator of stress associated with belonging to a minority group (‘minority stress’) that could attenuate benefits of volunteering (i.e., concealment of one’s SGM identity). This work was supported by an NIH Administrative Supplement to enhance recruitment and enrollment of diverse participants in aging research, specifically older SGM individuals.

Finally, given that the COVID-19 pandemic began in Year 3 (out of 5) of our study, we will also examine whether study participation pre- or post-COVID study modifications impacted findings.

## Methods

### Study design, setting, ethical considerations

This is a randomized trial registered at clinicaltrials.gov (NCT03343483). All procedures were approved by the Research Subjects Review Board at the University of Rochester. All subjects participate in study interventions for up to one year, with repeated assessments over the course of the study (baseline, 3, 6, 9, 12 months). Those randomly assigned to the volunteering condition receive the intervention at Lifespan as well as Lifespan volunteer placements in the community or at home (during COVID restrictions) per standard Lifespan policy and procedures. Those randomly assigned to the life review condition receive the intervention via the phone (for training) and their homes via email/mail.

### Participants

The study will recruit and enroll up to 375 community dwelling older adults (age 60 or older) to reach the target number of randomized subjects of n=150 in each group. Inclusion and exclusion criteria appear in Table 1, including any changes that were made in response to remote procedures for COVID precautions. Subjects are asked to refrain from initiating new long-term volunteer activities during their 12 months of involvement with the study. Mental health or physical health diagnoses or symptoms are not cause for inclusion/exclusion, but are assessed over the course of the study. Subjects are required to be able to read and write in English (per requirements of the study interventions).

**Table 1.**
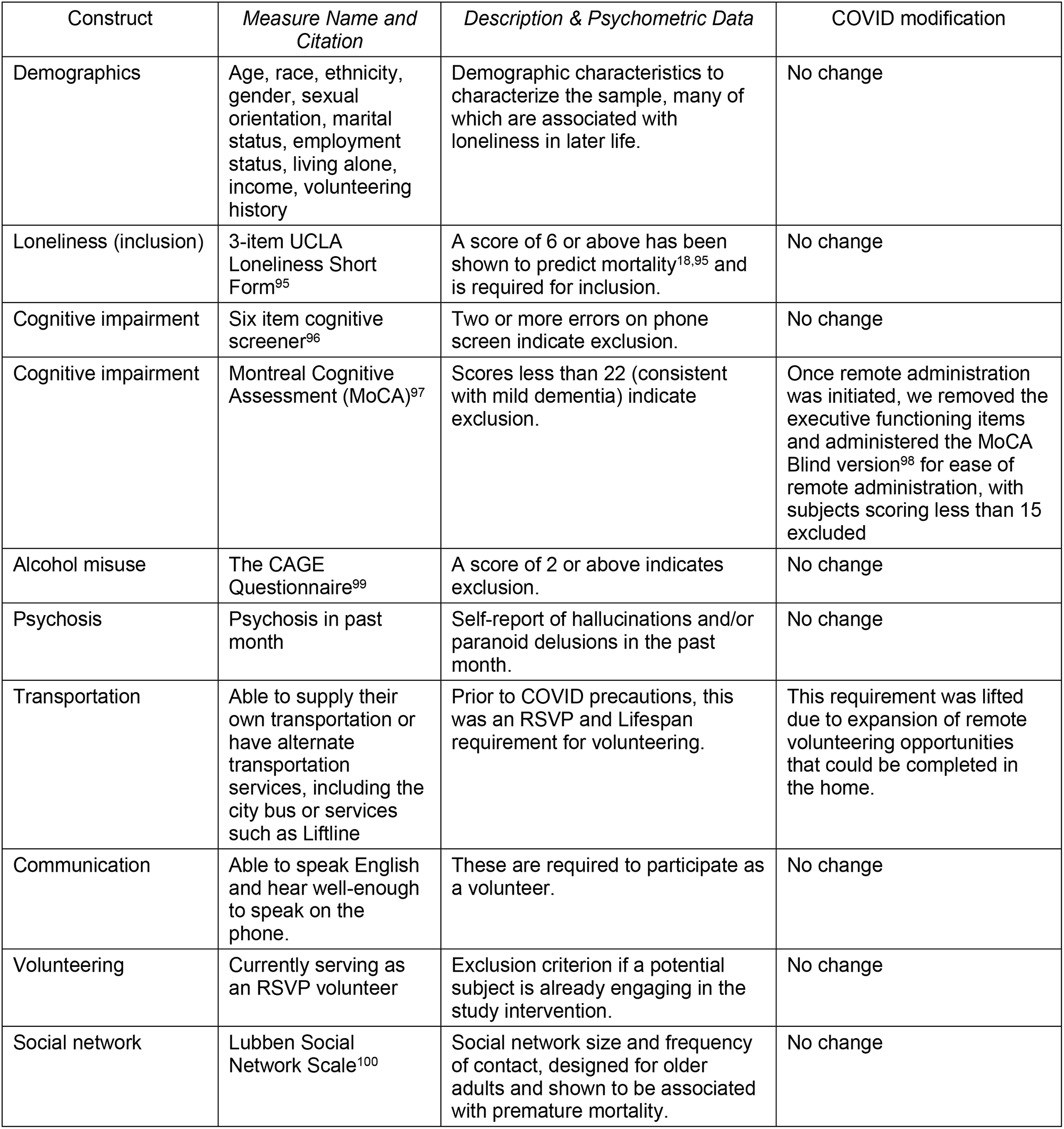
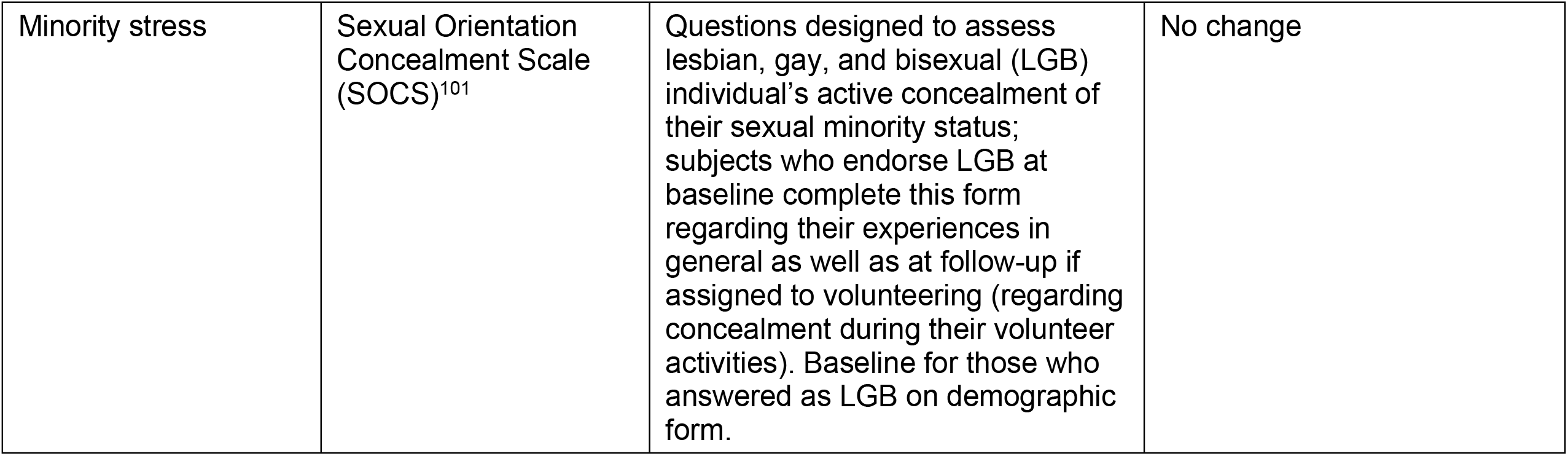
Baseline variables to assess inclusion/exclusion criteria and characterize the sample and modifications due to COVID-19

### Procedures

Our primary recruitment strategy is mailing information letters with a study brochure to adults 60 and older who received primary care within the UR Medicine healthcare system in the prior 2 years. Additional recruitment strategies include paid advertisements in local periodicals, flyers at various community sites, community presentations at senior centers and libraries, informational presentations to clinical providers likely to encounter lonely older adults (e., geriatricians, geriatric psychiatrists, neurologists) for direct referrals, and brochures distributed to Meals on Wheels clients. Potential subjects who contact study staff complete a brief phone screen to assess eligibility and provide additional information about the study purpose and activities. Those who are eligible and interested are invited to schedule a baseline interview in the PI’s laboratory at the University of Rochester Medical Center or via Zoom/phone (during COVID restrictions). The baseline interview includes a structured protocol for assessing capacity for informed consent, followed by assessment of additional exclusion criteria and baseline characteristics (see Figure 1, SPIRIT schedule of enrollment, interventions, and assessments).

**Figure 1:**
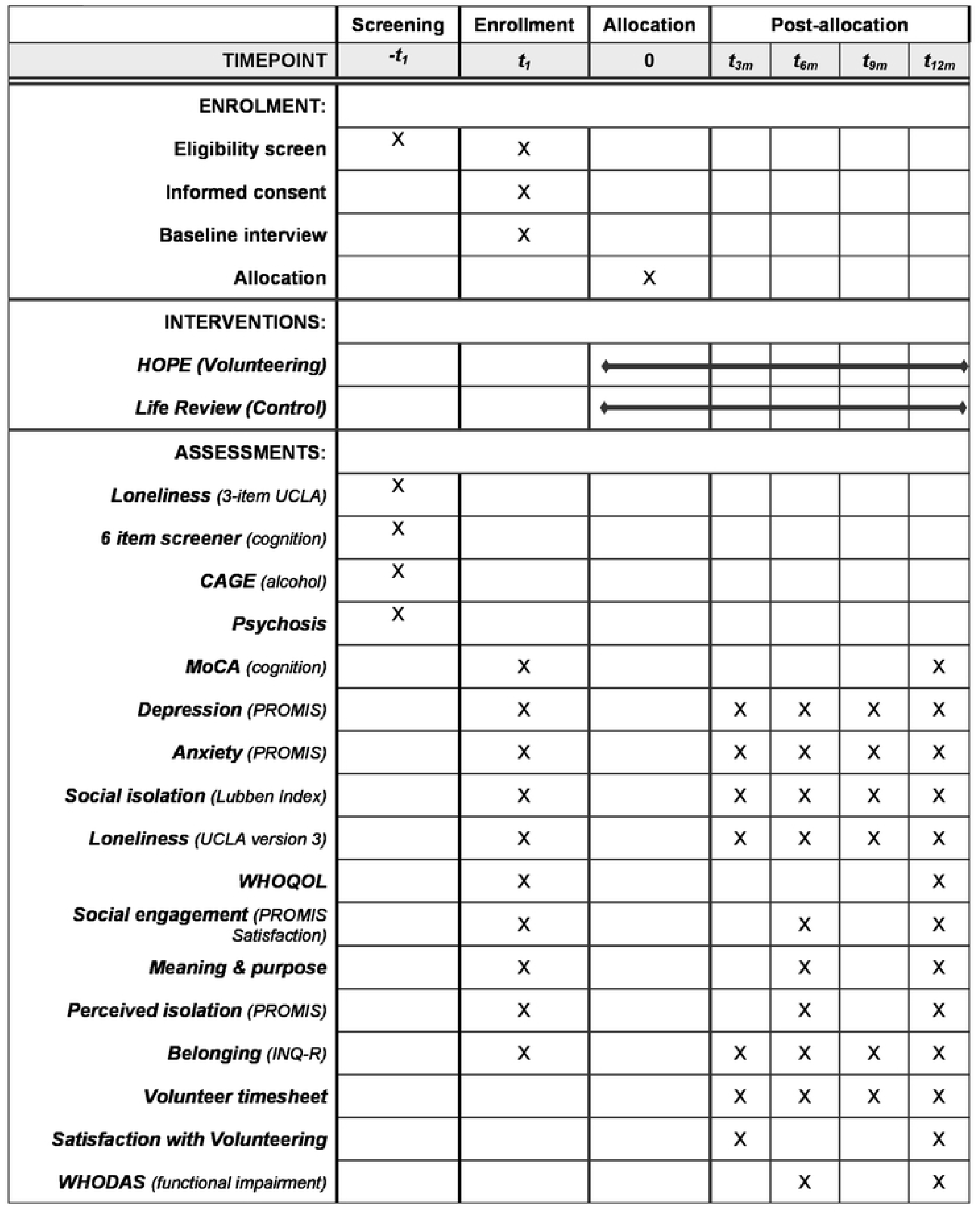
HOPE Study schedule of enrolment, interventions, and assessments (SPIRIT)

**Figure 2:**
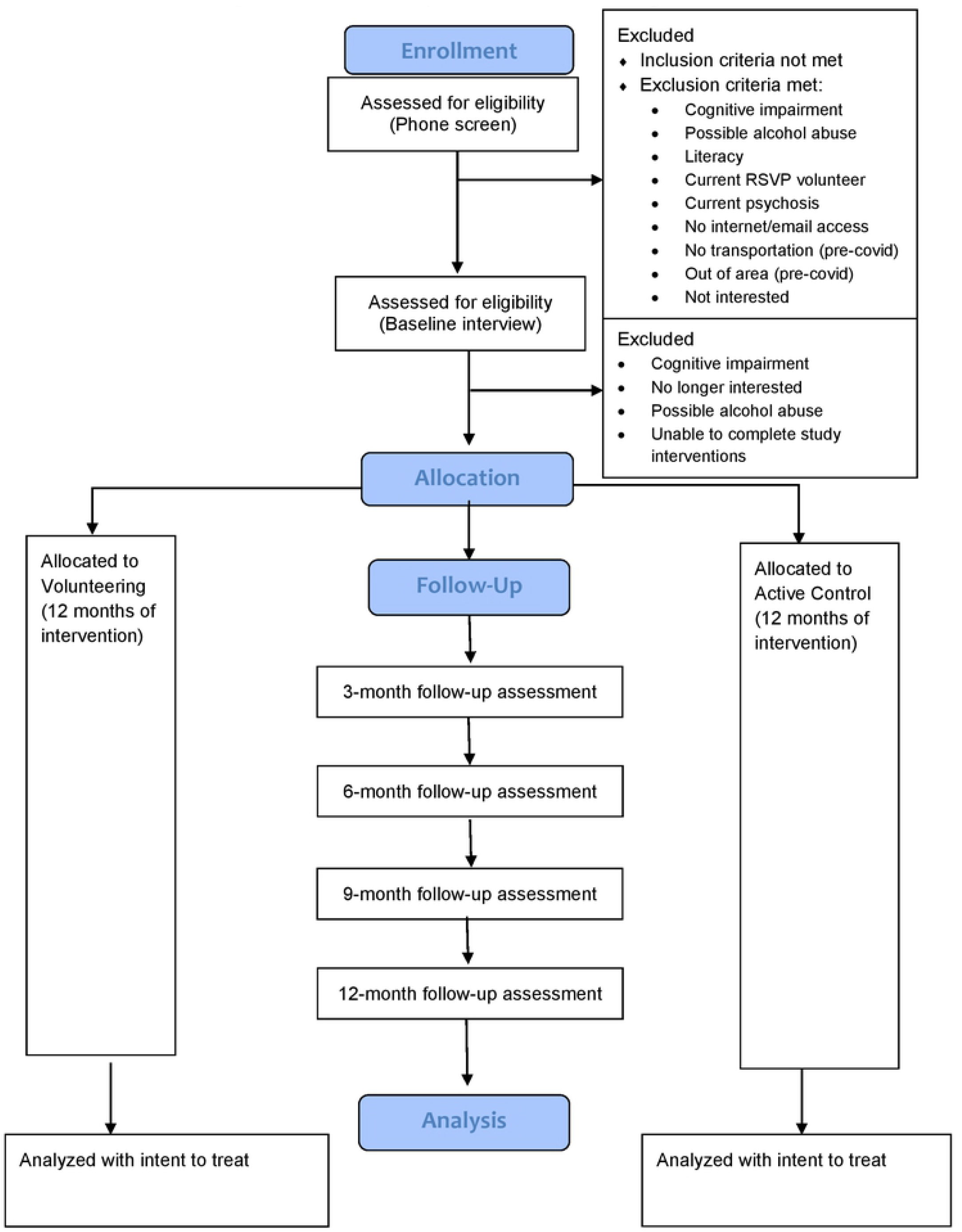
HOPE Study CONSORT Flow Diagram

### COVID-19 procedural modifications

Due to the COVID-19 pandemic, enrollment was paused in March 2020 until the University was allowed to resume research activities in April 2020. All study procedures were switched to entirely remote activities at that time. In-person study visits were changed to Zoom (HIPAA-compliant version) or phone. Study COVID restrictions have remained in place, with subject assessments remaining remote given that our subjects are older and thus more vulnerable to serious complications from COVID-19. While some potential subjects have declined participation due to the remote nature of assessments, that number is very small; study staff are available by phone to help subjects access email and learn to use Zoom.

### Randomization

Subjects are randomized (using the REDCap randomization module) to receive either the volunteering intervention or the control intervention. We used permuted blocked randomization with varying block lengths (unknown to PI and assessors). Neither assessors nor subjects are blind to condition. We have found that having the same assessor complete each assessment significantly improves retention. Our use of self-report measures as primary outcomes significantly reduces potential bias due to lack of blinding; our primary constructs, loneliness and quality of life, involve subjective perceptions, which are most directly measured by self-report measures.

### Volunteering intervention

Those subjects assigned to the HOPE intervention volunteer with the AmeriCorps Seniors RSVP program as part of their standard process for placing volunteers. The objective of RSVP is to match seniors with volunteer opportunities that match their interests and capabilities. Subjects begin training at Lifespan within two weeks of randomization. The target expectation for volunteering is 4 times per month (about once per week). Lifespan collects monthly timesheets, which include the number of hours and types of volunteer activities done. AmeriCorps Seniors provides a small reimbursement for travel to volunteers as needed for travel related to volunteering. The other component of the intervention is on-going training, volunteer support groups, and educational activities offered by Lifespan. These gatherings for volunteers serve to promote retention in the program, assist volunteers with any problems/issues that have arisen, and promote social connectedness among volunteers. The initial design for the study involved a single volunteer placement for all subjects that involved providing in-home companionship and respite care for older adults with dementia. However, acceptability of this volunteer placement was very low and potential subjects reported concerns about their capability to safely provide respite services for adults with more advanced dementia. In consultation with our community partner, Data and Safety Monitoring Committee, and with approval from our funder, the volunteering intervention was expanded to include the full range of volunteering placements available through RSVP, which significantly increased interest in the project. While some subjects do choose to provide respite for adults with dementia, there is a diverse range of other volunteer opportunities, including volunteering at animal shelters, delivering for Meals on Wheels, helping at a food bank, driving older Veterans to appointments, mentoring and tutoring youth, and helping run educational and wellness programs (e.g., Tai Chi, Matter of Balance). The program also creates new placements for volunteers based on interests and capabilities. All placements are documented by Lifespan and research staff. To establish new RSVP placements, the volunteer coordinator establishes a Memorandum of Understanding regarding information sharing to best support volunteers (and act as a liaison and ombudsman if needed) and to document hours served for reporting purposes (and mileage reimbursement).

### Volunteering intervention COVID-19 modifications

Once study procedures switched to entirely remote in Year 3 of the trial, volunteering activities and placements were altered to allow only activities completed in the home, such as friendly calling to other older adults, including those in long-term care settings experiencing significant isolation. In June 2020, the University IRB approved volunteering options returning to the community in addition to at home opportunities, when such opportunities were deemed safe by our community partner. Placements in the community are reviewed by RSVP and Lifespan staff and must have a documented COVID-19 safety plan in place before volunteers engage in volunteer opportunities. Before resuming these in-person activities, we consulted our Data Safety Monitoring Committee for consultation and approval. At the time of writing this paper, subjects are allowed to volunteer in the community if they prefer, or continue volunteering activities that can be completed at home (e.g., friendly calling) in line with RSVP practices of personalizing the volunteer experience to match preferences. Other safe remote options have included delivery of meals for Meals on Wheels (using physical distancing procedures) and projects for Lifespan and other agencies that involve graphic design, photography, or writing.

### Control condition

Those subjects assigned to life review complete a series of self-guided (with email/postal mail support) life review writing exercises over 12 months. The active control condition was chosen to control for (and minimize confounding by) potential non-specific effects of participating in a study intervention; expectancies about benefit; and starting a new cognitively engaging activity. This standardized and evidence based intervention is commonly used to reduce depression and promote well-being in later life; further, it was validated as a self-guided intervention to improve well-being by Lamers and colleagues.^80^ To minimize the social nature of providing the intervention (i.e., minimizing confounding our conditions), the life review is largely self-guided, including replacing the counselor (and one-on-one sessions) with email/postal mail support and a self-help book, per the protocol of Lamers and colleagues.^80^ Subjects complete two sections of the life review (with the self-help book) each month and send ‘assignments’ twice per month to the ‘Life Review Coach’ who responds with supportive comments.

### Study variables

Outcome variables are described in Table 2. Our study has two primary outcomes, loneliness and quality of life. Loneliness is assessed at all time points (baseline, 3 months, 6 months, 9 months, and 12 months), while quality of life is assessed only at baseline and 12-months (as it is a lengthier assessment). Two secondary outcomes assess potential mechanisms whereby volunteering may reduce loneliness (social engagement and meaning/purpose in life). Given that few RCTs have been conducted with older adults to examine loneliness and there is a limited literature to guide selection of measures of social connection that are most sensitive to change, we also included two additional secondary outcomes that are similar to, but not redundant with, the UCLA Loneliness Scale—belonging (Interpersonal Needs Questionnaire) and perceived social isolation (PROMIS). Moderator variables include ‘dose’ of volunteering and satisfaction with volunteering, as well as functional impairment and proportion of study participation completed after COVID modifications were put in place. For exploratory analyses with sexual and gender minority identity (SGM; e.g., lesbian, gay, bisexual, transgender), we assess SGM identity as well as one indicator of minority stress—concealment of one’s SGM identity (Table 1). Finally, for all subjects, we are conducting qualitative interviews to obtain perceptions of the benefits (and/or potential harms) of study interventions, including any impact aspects of subjects’ identities may have had on their experiences (e.g., race, gender, sexuality, etc.).

**Table 2.** Study variables

|  |  | *Measure Name and Citation* | *Description & Psychometric Data* |
| --- | --- | --- | --- |
| Loneliness | Primary | UCLA Loneliness Scale, version 3<sup>102</sup> | Self-report (20 items). Yields a continuous total score, with greater scores indicating greater loneliness (range 20 – 80). None of the items use the word 'lonely' to reduce under-reporting due to social desirability and stigma. It has demonstrated strong psychometric properties with older adult samples.<sup>13,102</sup> |
| Quality of life | Primary | World Health Organization Brief Quality of Life Scale (WHOQOL-Bref).<sup>103</sup> | Self-report (26 items). Yields a continuous total score, with greater scores indicating greater quality of life (range 0-100). Scores for four domains (physical, psychological, social, and environmental) are also available; social and emotional quality of life are expected to improve most in our study. It has demonstrated strong psychometric properties.<sup>104</sup> |
| Social engagement | Secondary (mechanism) | PROMIS Satisfaction with Social Roles and Activities<sup>105</sup> | This computerized adaptive test (CAT) produces T scores with a mean of 50 and standard deviation of 10. Greater scores indicate greater satisfaction with social roles and activities. |
| Meaning & purpose | Secondary (mechanism) | Patient-Reported Outcomes Measurement Information System (PROMIS) Meaning and Purpose | This computerized adaptive test (CAT) produces T scores with a mean of 50 and standard deviation of 10. Greater scores indicate greater meaning and purpose. |
| Perceived social isolation | Secondary | PROMIS Social Isolation computerized adaptive test<sup>106</sup> | This computerized adaptive test (CAT) produces T scores with a mean of 50 and standard deviation of 10. Greater scores indicate greater isolation. |
| Belonging | Secondary | Interpersonal Needs Questionnaire, belonging subscale<sup>107,108</sup> | Self-report (9 items). Yields a continuous total score, with greater scores indicating greater belonging (range 0 – 18). It has demonstrated strong psychometric properties with older adults. |
| Volunteering quantity | Moderator | RSVP Volunteer Timesheet | Collected by the volunteer coordinator, this standard reporting form documents amount of time spent volunteering each month. |
| Volunteering quality | Moderator | Satisfaction with Volunteering Scale | This is a standard AmeriCorps Seniors program evaluation survey. |
| Functional impairment | Moderator | World Health Organization Disability Assessment Schedule (WHODAS) total score.<sup>109</sup> | Functional impairment across several domains, mobility, cognition, self-care, social, emotional. |
| Participation during COVID | Moderator | Proportion of study participation completed after COVID modifications were in place. | Calculated by randomization date and COVID procedural modifications start date of April, 2020. |
| Experiences in study interventions | Exploratory | Qualitative interview | Open-ended feedback on study interventions. |

### Sample size calculation

A power analysis was conducted to test treatment effects by the intent-to-treat (ITT) analysis for the primary outcomes. We assumed a conservative 20% attrition rate and a 0.3 within-subject correlation. A sample size of N = 300 (or N = 150 per treatment group) will allow us to detect a small effect size of 0.2 for loneliness, with 80% power based on a two-sided type I alpha = 0.05. The assumed within-subject correlation of 0.3 reflects the relatively long time lag between consecutive assessments. For Aim 2 examining mechanisms, the proposed sample size also has 80% power to detect 32% mediation effects for loneliness, a continuous outcome. If full mediation is not achieved for any of the mediators considered, 32% mediation effects are sufficiently large to be of clinical importance for the outcomes of interest.

## Data Analytic Strategy

Descriptive statistics will summarize distributions of each outcome, with means and standard deviations for continuous outcomes and percent for categorical outcomes. Two-sample t tests (or the Mann-Whitney-Wilcoxon rank sum test) and Chi-square tests will be used to examine balance of treatment randomization for continuous (if distributions are highly skewed, highly skewed, especially with outliers) and categorical variables. We will examine the potential impact of COVID-19-related study modifications by computing the proportion of intervention time after COVID modifications were implemented.

Aim 1, the effect of volunteering on the co-primary outcomes of loneliness and quality of life (both continuous variables), will be tested using semiparametric weighted generalized estimating equations (WGEE) for longitudinal regression analysis. WGEE imposes no analytic model for the distribution of the response (dependent variable) and thus provides valid inference regardless of how the response variable is distributed. Moreover, it provides valid inference under the missing at random (MAR) mechanism, the most common in clinical research studies, if the missing data is correctly modeled.^81^ These models use an Intent to Treat design, with all subjects randomized to either condition included in the analysis, regardless of their compliance to their assigned interventions, to ensure replicability of treatment effects in similar study populations. For Hypothesis 1a, loneliness will be the response variable, with condition, time and their interaction as the predictors, controlling for age and gender. We hypothesize that there will be an effect of condition on loneliness at all follow-up points indicating differing levels of loneliness in the direction: control > volunteering. If a significant difference exists (a significant time by condition interaction), appropriate linear contrasts will confirm the hypothesized directional effects (greater loneliness for the control group). For Hypothesis 1b, the same analytic strategy will be used, but with quality of life as the response variable and anticipating greater quality of life in the volunteering condition. For both models, we will include a time-varying indictor to examine whether the intervention time once COVID modifications were implemented is associated with loneliness and quality of life. Moderation analyses for Aim 3 (functional impairment) will be conducted using the same analytic strategy.

Aim 2 examines mechanisms (mediation) whereby volunteering reduces loneliness and improves quality of life. This aim will be examined by structural equation models (SEM)^82 83^ to test the putative mediators of purpose in life and social engagement. If the outcome and mediator have highly skewed distributions, we will use semiparametric methods for more robust inference in these models as well as apply appropriate variable transformation to improve efficiency.^83,84^ We will report standard goodness-of-fit measures—chi-square test, the comparative fit index (CFI), the index of Tucker and Lewis (TLI), and Root Mean Square Error of Approximation (RMSEA).^82,85,86^ Increases in social engagement (PROMIS Satisfaction with Social Roles & Activities) at 6 months is hypothesized to mediate the effect of intervention condition on decreased loneliness at 12 months. The SEM-based mediation models will be applied to test the hypothesis, with social engagement as the mediator, intervention condition as the predictor and loneliness as the outcome, controlling for age and gender. If the null of full mediation is rejected, we will estimate direct, indirect and total effects to assess the strength of mediation. The same analytic strategy will be used for Purpose in Life.

Aim 3 conditions under which volunteering may provide maximal benefit, including ‘dose’ (greater hours volunteering) and greater satisfaction with the intervention. Dose-response relationships (Aim 3) will be examined by structural mean models to supplement the ITT analysis for Aim 1, which provides intervention effects averaged over all subjects randomized to the intervention conditions. When intervention compliance (volunteering hours/satisfaction for subjects in the volunteering condition) demonstrates a dose-response relationship, as we hypothesize in Aim 3, complier average causal effects (CACE) will model and test such a dose-response relationship. CACE is a compliment to ITT analyses, which answers the question whether the intervention has any treatment effect for the study population as a whole regardless of compliance, rather than the question whether the intervention has any therapeutic value and if so, how the therapeutic effect changes with increased dose.^87^ CACE provides different intervention effects for individuals depending on their levels of compliance, which can be quite informative, especially when there is large variability in intervention compliance and strong dose-response relationships. Since volunteering time is only required for the volunteering condition, standard statistical models cannot be used to perform CACE analysis. The CACE approach enables an estimate of the treatment effect at each level of “compliance” (i.e., amount of hours volunteered), without the need for a measure of compliance in the control group. In this way, we will be able to tell how well volunteering reduces loneliness at different “doses” of volunteering. If results regarding study participation during the pandemic suggest differential efficacy as a function of participation once COVID-related study modifications were put in place, we will also conduct a CACE analysis accordingly to examine ‘dose’ of volunteering pre-COVID (i.e., before volunteer opportunities were restricted). Given that the control condition was unchanged during the pandemic, this analysis may be best to detect potential effects of COVID-19 on study outcomes.

We will use the latest semi-parametric structural mean model (SMM)^88-90^ based on the structural functional response models (SFRM) for our CACE analysis, which not only allows for continuous, but also multiple dose variables. ^88-90^ We are particularly interested in potential non-linear dose-response relationships so that we may determine optimal dose intervention whereby increased exposure (i.e., number of hours volunteered) becomes less worthwhile (in terms of reducing loneliness). We will apply the SFRM-based SMM to analyze dose-response relationships. We will first model dose using non-parametric methods such as LOWESS curves and then characterize the patterns using parametric methods for inference and improved efficiency. This allows us to capture detailed dose and response relationships and provide more interpretable findings.

**Data and safety monitoring** for this study is overseen by a Data and Safety Monitoring Committee composed of individuals independent of the study as well as the PI and one Co-I; none are at a different institution given the low-risk nature of the study intervention; selection of a non-clinical population; and study design without blinding. Per the NIH Data Sharing Policy, de-identified data will be available to interested investigators under a data use agreement.

### Timeline

Enrollment for this trial is underway and expected to finish by May 2022, with completion of follow-up assessments through April 2023, and completion of primary outcomes soon thereafter.

## Discussion

The outcomes of this study will have several implications. The numerous negative health outcomes associated with loneliness in older people have rendered loneliness itself a new public health target.^119^ The COVID-19 pandemic and the need for physical distancing introduced loneliness into everyday conversation and powerfully demonstrated the deleterious effect of loneliness on health and well-being. However, healthcare in the U.S has not capitalized on social connection as preventive medicine. Unlike other countries where social and health services are more fully integrated, such as the UK’s national strategy for addressing loneliness,^91^ in the U.S., fee-for-service payment models and the separate funding and functioning of health and human services disincentivize provision of non-medical services by health care providers and systems. The COVID pandemic further stretched the health care sector, making attention to social determinants less feasible. However, the pandemic also made clear that loneliness and social isolation contribute powerfully to illness as well as utilization and cost of health services. Raised awareness of the adverse health consequences of loneliness may reinvigorate efforts to address the problem as a component of integrated health care.^92^

In order to do so, the evidence-base on interventions for loneliness, which is limited currently and does not clearly indicate which interventions are most effective, must be improved. This Stage III efficacy trial (NIH Stage Model) is designed to examine efficacy with the intervention provided as it is in the community, nationwide, to reduce the time from testing to dissemination and implementation with older adults who might benefit. This study design has both strengths and limitations that will impact interpretations of results and next steps. The need to tailor the volunteering activity to individual preferences is the foundation of the national RSVP model and was needed to promote acceptability of the project; while a single volunteer placement for all volunteers would have reduced heterogeneity in the intervention, such a restriction was both not feasible and not representative of volunteering programs available in the community. The RCT design increases internal validity, but introduces other challenges, including the fact that participating in a research study is itself a volunteer opportunity that provides ongoing social contact (potentially confounding conditions and attenuating effects). Our study was also impacted by the COVID-19 pandemic, including a pause in enrollment (though relatively brief, only two months), as well as changes in the types and variety of volunteering placements offered.

Given that our study was not designed to specifically study loneliness in SGM older adults, our study interventions were not tailored to this group; this includes a lack of SGM-specific volunteer opportunities (e.g., volunteering for SGM community organizations), which is a feature of volunteering shown to increase SGM community connectedness,^79^ which in turn, may be an effective strategy to reduce loneliness.^93^ Without this increase in SGM community connectedness, SGM volunteers may not experience the hypothesized benefits of volunteering. We will also examine the role of concealment (i.e., hiding one’s identity from others) while volunteering because this aspect of minority stress may play a role in both social connection and health outcomes among SGM older adults.^94^ Participants who engage in concealment while completing their volunteer service could experience an *increase* in loneliness; if this is the case, SGM-specific volunteer activities would be advisable.

Older adults who report loneliness are less likely to actively seek out volunteering opportunities; if our results support efficacy of volunteering for reducing loneliness and improving quality of life for older adults experiencing loneliness, dissemination and scaling up efforts may involve connecting primary care patients who are lonely with The Senior Corps through aging services agencies, which we have shown to be feasible in our companion study, The Senior Connection.^97^ National infrastructure for the volunteering program for older adults tested in our study—AmeriCorps Seniors—ensures that volunteering is highly scalable. Existing infrastructure will make it possible to engage a large proportion of lonely older adults in volunteering. Future directions for research on the benefits of volunteering for social health and well-being include examining cultural and other psychosocial determinants of loneliness that may impact willingness or ability to engage in volunteering (e.g., history of psychological trauma), or that may require tailoring of activities (e.g., under-represented backgrounds or disabilities). Future work should also consider strategies to increase engagement in volunteering programs by older adults not likely to seek out such activities on their own, as well as the potential health impacts of reducing loneliness to provide information on utilization and cost that is needed to support adoption of care and payment models that integrate assessment and treatment of loneliness into health care.

## Data Availability

This protocol paper does not present results.

## Authors’ contributions

KVO was the principal investigator and obtained funding. KVO, YC, XT, BC, SS, and GW designed the study. KVO oversaw its implementation and supervised the research assessments. JR directed the volunteering program at Lifespan and DP served as the volunteer coordinator and supervised volunteers and coordinated data collection on volunteer hours. AS and KVO developed the life review control condition materials. AB, EB, and AS helped with study set up and administration, development and implementation of COVID modifications, and development of recruitment and enrollment procedures. AB oversaw coordination with our community partner, Lifespan. In addition to KVO (first author), AB, AS, AVB, BC, XT, and YC were involved in writing the text. All authors reviewed and approved of the paper before submission.

## Acknowledgments

The study was supported by a grant from the National Institute on Aging (R01AG054457, R01AG054457-02S1, Van Orden, PI). We wish to thank all members of the study team, the staff of Lifespan, the many agencies and clinicians who assisted in developing and implementing recruitment plans, and our study participants who generously gave their time and effort to study programs that can improve quality of life for older adults.

---

Figure 1. SPIRIT Schedule of enrollment, interventions, and assessments (separate attachment)

Figure 2. CONSORT Diagram (separate attachment)

Figure 3. SPIRIT Checklist and protocol (separate attachment)

